# Covid-19 vaccination decisions and impacts of vaccine mandates: A cross sectional survey of healthcare workers in British Columbia, Canada

**DOI:** 10.1101/2024.12.09.24318733

**Authors:** Claudia Chaufan, Natalie Hemsing, Rachael Moncrieffe

## Abstract

**Background:** Since vaccination mandates in the healthcare sector were introduced across Canada, public health authorities in the province of British Columbia implemented among the strongest ones in the country. While some workers unions opposed these mandates, they were supported by most health establishments, policymakers, and academics. Ensuing labour shortages compounded the ongoing health crisis in the province, leading to mounting calls to lift mandates and allow non-compliant, terminated or suspended, workers to return and ease pressures on the system. Nevertheless, mandates remained effective until July 2024, over one year after the World Health Organization had ended its declared Covid-19 global health emergency. Most research has focused on the perceived problem of vaccine hesitancy among healthcare workers, yet not on their lived experience of the policy or their views on its impact on access to, and quality of, patient care.

**Goal:** To document the experience and views on mandated vaccination of healthcare workers in British Columbia.

**Methods:** Between May and July of 2024, we conducted a cross-sectional survey of healthcare workers in British Columbia. We recruited participants through a snowball sampling approach, including professional contacts, social media, and word-of-mouth.

**Results:** Close to half of respondents, with 16 or more years of professional experience, were unvaccinated, and most had been terminated due to non-compliance with mandates. As well, and regardless of vaccination status, most respondents reported safety concerns with vaccination and felt unfree to make their own vaccination choices, yet did not request exemptions due to high rejection rates by employers. Most of them also reported experiencing anxiety or depression, with about one fourth considering suicide, as a result of mandates. Nevertheless, most unvaccinated workers reported satisfaction with their choices, although they also reported significant, negative impacts of the policy on their finances, their mental health, their social and personal relationships, and to a lesser degree, their physical health. In contrast, within the minority of vaccinated respondents, most reported being dissatisfied with their vaccination decisions, as well as having experienced mild to serious post vaccine adverse events, with over half within this group reporting having been coerced into taking further doses, under threat of termination, despite these events. Further, a large minority of all respondents reported having witnessed underreporting or dismissal by hospital management of adverse events post vaccination among patients, worse treatment of unvaccinated patients, and concerning changes in practice protocols. Nearly half also reported their intention to leave the healthcare industry.

**Discussion:** Our findings indicate that in British Columbia, mandated vaccination in the healthcare sector had an overall negative impact on the well-being of the labour force, on the sustainability of the health system, on patient care, and on ethical healthcare practice. Findings resemble those of a similar study in the province of Ontario, with perhaps the most salient difference being that in British Columbia the policy was implemented at the provincial, rather than the healthcare establishment, level, leaving no room for individual establishments to opt out.

**Conclusions:** Measured against the 2021 criteria proposed by the Organization for Economic Cooperation and Development to evaluate the merits or lack thereof of public policy, the policy of mandated vaccination in British Columbia failed on several fronts - scientific, pragmatic, and ethical. Future research should examine why this and similar policies persist despite the evidence against them. Findings from this and similar studies should be considered, especially during emergencies, to guarantee the quality of the evidence informing policy, health systems sustainability, and the human rights to bodily autonomy and informed consent of both healthcare workers and members of the public.

## Introduction

On August 12, 2021, Dr. Bonnie Henry, the British Columbia (BC) Provincial Health Officer (PHO), announced the *Residential Care Covid-19 Vaccination Status Information and Preventive Measures* order. This order required that healthcare workers (HCWs) in long term care and assisted living facilities provide proof of completing a primary series of Covid-19 vaccination by October 12, 2021 or face termination (BC Ministry of Health, 2021; Office of the Provincial Health Officer, 2022c). On September 12, 2021, the *Hospital and Community COVID-19 Vaccination Status Information and Preventive* Measures order was introduced. This second order required that HCWs in the provincial health authority and five regional health authorities in BC, provincial mental health facilities, and hospital and community care settings - including employees, contractors, volunteers and students, administrative and fully remote workers - provide proof of vaccination by October 26, 2021 to maintain employment (Office of the Provincial Health Officer, 2022b). The Public Health Officer (PHO) Dr.

Henry stated that because of the “risk inherent in accommodating persons who are not vaccinated,” only medical exemptions would be approved, and only “on the basis that vaccination would so seriously jeopardize the individual’s health that the risk to the individual’s health posed by vaccination outweighs the benefit” (Office of the Provincial Health Officer, 2022b) (page 27). Dr Henry claimed that the measures reflected a “balancing the interests of the people working or providing services in the hospital and community care sectors… against the risk of harm posed by unvaccinated people working or providing services in the hospital or community care sectors” (Office of the Provincial Health Officer, 2022b) (page 8).

In contrast to the strong support for vaccination mandates from most healthcare organizations, some unions, such as the BC Nurses Union (BCNU) and the Hospital Employee Union (HEU) expressed concerns and stated their preference for a voluntary vaccination program given the risks posed by mandates to an already understaffed healthcare system (BC Nurses Union, 2021b; Hospital Employee Union, 2021; Slepian, 2021). Like other provinces in Canada, there is a healthcare crisis in BC, specifically a critical shortage of nurses and family doctors (Ahmed & Bourgeault, 2022; BC Nurses Union, 2021a), exacerbated by population growth, a shrinking healthcare labour force, and an ongoing toxic drug crisis in the province. The BCNU president noted that nurses in BC had long cautioned about a staffing shortage, noting that there were over 5,300 nurse vacancies as of 2023, and that by 2031 it is projected that an additional 27,000 nurses will be needed to meet the demands of a growing population (BC Nurses Union, 2023). Therefore, in a media statement on September 16^th^, 2021, the BCNU stated that it “cannot support any order which will serve to remove even a single nurse or [HCW] from the healthcare system at a time of severe crisis”. The union cautioned that, while encouraging vaccination among their members, “taking nurses away from the bedside will have serious impacts on patient care”, and expected “government and health employers to avoid any measures that may take nurses away from providing patient care” (Slepian, 2021). Further, the BCNU expressed concerns about the negative impact of mandates on worksites already experiencing labour shortages (BC Nurses Union, 2021b), and explained that “in some rural communities, losing just a single nurse or health-care worker from the system” can be “disastrous” (Shaw, 2023).

Opposing views notwithstanding, vaccine mandates for HCWs have generally received widespread support, not only by health establishments and officials, but also in the peer-reviewed academic literature, especially in relation to the perceived role of HCWs as “trusted sources” of vaccine information (Achat et al., 2022; Dietrich et al., 2022; Evans et al., 2022). Therefore, “vaccine hesitancy” has been, and continues to be, framed as a problem to be overcome, with greater vaccination uptake presented as the undisputed, desired outcome, ideally achieved through information and education, albeit with mandatory vaccination when necessary (Dietrich et al., 2022; Lee et al., 2022; Oberleitner et al., 2022). For example, in a study evaluating the impact of the mandate in BC, the authors concluded that the mandate was successful because it increased vaccination uptake among HCWs in BC (Okpani et al., 2024). However, there has been limited exploration of HCWs’ own experiences and perspectives on vaccination mandates, without pre-existing assumptions about the desirability of vaccination. To help fill this gap, we conducted a survey in BC, Canada, to gather HCWs’ views on workplace vaccination mandates and on how the policy affects the capacity and quality of the healthcare system.

## Background

Vaccination mandates, especially among HCWs, has been a contentious issue, especially in the Covid era. Since the outset, government and public health officials have deemed that accommodating even small proportions of unvaccinated HCWs was too great of a risk for patients, the public, and the fragile healthcare system. When the mandate was first announced in BC, levels of vaccination among HCWs were already high. As per Health Minister Adrian Dix’s own admission during an unrelated press conference, and in response to concerns regarding healthcare labour shortages, “the massive majority of [HCWs] have been immunized everywhere in B.C.”; Dix further explained that vaccination rates among staff were likely to be even higher than the already high rates (87%) among the general population of adults in the province (Slepian, 2021). Nevertheless, the Health Minister ultimately insisted that vaccination mandates for HCWs were essential to keep patients safe and end the pandemic (Slepian, 2021). These comments echoed Dr. Henry’s order, which asserted that “any slippage in the level of vaccination in the health-care workforce” could undermine healthcare system capacity (page 3).

Overall, supporters of the vaccination mandate – primarily healthcare organizations such as the Doctors of BC, the BC Dental Association, and the College of Pharmacists of BC, among others (BC Chiropractic Association, 2021; BC Dental Association, 2021; Doctors of BC, 2021) – have emphasized the benefits of the policy for the public good, while minimizing or dismissing the risks. Salient cases include the president of Doctors of B.C., Dr Mathew Chow, who affirmed his support for a mandatory vaccination policy (Doctors of BC, 2021) “in the health care sector and beyond” and further stated that “in the present circumstances and with very few exceptions, full participation in society will require vaccination (Doctors of BC, 2021). Similarly, the Board of Directors of the BC Chiropractic Association, issued a statement expressing strong support for Covid-19 vaccinations and for mandates for HCWs, criticizing the views of members who opposed the vaccines due to safety concerns as a “misrepresentation of the facts” (BC Chiropractic Association, 2021).

Ultimately, an estimated 2,500 HCWs in the province were terminated for failing to comply, in whole or in part, with Covid-19 vaccination mandates, with over half of them employed in Interior and Northern Health, regions where labour shortages have resulted in ongoing emergency room closures (DeRosa, 2023). While provincial government and public health officials have minimized the impact and risks of imposing mandates on labour shortages (Burns, 2023), over time, and with the ongoing healthcare crisis in the province, there have been mounting calls for mandates to be lifted so that laid-off or suspended HCWs could return to work and ease pressures in the system. These have included, for example, calls from members of the Union of BC Municipalities (Union of BC Municipalities, 2023), the provincial government opposition (Shaw, 2023), and several mayors and elected Members of the Legislative Assembly (MLAs) (DeRosa, 2023), who have repeatedly urged the government to lift the vaccination mandate for HCWs. For example, the mayor of a rural community objected to BC “being the only province left in Canada still on this stand”, while their hospital faced repeated closures due to a shortage of nurses and doctors (Shaw, 2023). The MLA for another rural area, Peace River North, Don Davies, argued that "As our public health care system wavers on the utter brink of collapse, the exclusion of these professionals not only undermines our healthcare capacity but also erodes public trust in our healthcare" (Cunha, 2024).

The government and public health officials also faced criticism for inconsistent expectations for different categories of healthcare professionals. While the PHO announced in March 2022 that unvaccinated HCWs from 18 colleges (including nurses, midwives, psychologists, optometrists, physical therapists, traditional Chinese medicine practitioners, and dental hygienists) would be barred from practicing, they ultimately decided against this order, and opted instead for an “informed consent system” that required implicated HCWs to report their vaccination status to their colleges (Daflos & Weichel, 2022; Office of the Provincial Health Officer, 2022a). In response, both the BCNU and the Ambulance Paramedics and Dispatchers of BC expressed dismay over the inconsistent messaging and differential treatment of HCWs, arguing that terminated members should be reinstated (Daflos & Weichel, 2022). Be that as it may, BC was the last province in Canada to maintain emergency powers and vaccination mandates for HCWs. The order remained in place until July 26, 2024, when it was finally rescinded, and the PHO declared an end to the public health emergency and associated emergency powers in the Public Health Act (BC Ministry of Health, 2024). Notably, this rescission came over one year after the World Health Organization declared, in May 2023, that Covid-19 was no longer a global health emergency (World Health Organization, 2023).

## Methods

We conducted an online survey of British Columbia HCWs. Eligibility criteria included anyone working in a healthcare setting – whether in patient care, administrative, or maintenance services – in the province, and of any age and vaccination status. We used a snowball sampling method whereby respondents reached via professional networks of the lead author were invited to disseminate recruitment materials through social media and among their own networks, at seven-day intervals over one month (McDonald et al., 2024). Of the 372,792 people estimated to be employed in British Columbia in healthcare and social assistance in 2023 (Work BC, 2023), we recruited a convenience sample of 166 HCWs.

We pilot tested the survey with a local medical doctor and upon integrating their feedback, we launched it in May 2024, collecting responses through July 2024. The survey consisted of 15 sections, with sections automatically skipped depending on vaccination status or job termination for non-compliance with mandatory vaccination. After informing respondents about the purpose of the research and the confidentiality of the data, confirming that they worked in British Columbia, and obtaining informed consent, they were asked about their employment status and history, their experience of making vaccination decisions, the impact of the policy on their finances, personal and social relations, and mental and physical health, and their perspectives on the impact of the policy on patient care.

The survey consisted of 91 questions, including multiple choice, short answer, and Likert scale (i.e., rank ordered responses). Sections included: demographics (8 questions); employment (5 questions); Covid-19 experiences (2 questions); informed consent (11 questions); vaccination decision making (4 questions); vaccine side effects (3 questions); accommodations (3 questions); personal impact of vaccination policies (9 questions); self -rated health changes (4 questions); vaccination requirements and employment status (2 questions); impacts of job termination (10 questions); impacts on patient care (24 questions); experiences of administering Covid-19 vaccines (5 questions); and an open ended question for further comments (1 optional question). Respondents were entered into a raffle for a $100 gift card.

### Ethical considerations

The study was conducted following the Declaration of Helsinki (as revised in 2013). The study was approved by the York University Office of Research Ethics (No. 2023-389). Potential study participants were provided an information letter and consent form, including details on the study aims, methods, potential benefits and risks, and information about confidentiality and consent. They were informed of their right to withdraw consent at any time without consequences. The online survey questions were only accessible to participants after providing their informed consent.

### Statistical analysis

Due to the exploratory nature of the study, we performed only a descriptive analysis of the data using Excel spreadsheets. Authors met regularly to review the data, discuss the analysis, and identify trends.

## Findings

### Demographics

Most respondents (101/166, 61%) were between the ages of 45-54 years (55/166, 33.1%) and 55-64 years (46/166, 28%), followed by ages 35-44 years (32/166, 19.3%). Most respondents were women (112/166, 67.5%), middle-income (87/166, 52.4%), born in Canada (123/166, 74.1%), Caucasian/White (135/166, 81.3%), and married or living with a partner (107/166, 65%). A large minority (69/166, 42%) reported no caretaking responsibilities, whereas another large although slightly smaller minority (62/166, 37.3%) reported caretaking responsibilities of children or stepchildren, and a smaller proportion (26/166, 16%) had caretaking responsibilities with parents (Table 1). The most reported profession/area of occupation was nursing (25/166, 15.1%). About one-third of respondents (55/166, 33.1%) reported between 6 and 15 years of experience in their most recent career, close to one-fourth (43/166, 26%) between 16 and 25 years of experience, over one-fifth (39/166, 23.5%) over 26 years of experience, and a minority (19/166, 11.4%) five or fewer years. A large minority (68/166, 47%) reported 10+ years of education/training, followed by close to one- third between 0-4 years (50/166, 30.1%), and over one-fifth (38/166, 23%) reporting 5-9 years of training. About one third of respondents (49/166, 29.5%) reported working in the Interior Health region, followed by Fraser Health (40/166, 24.1%) and by Vancouver Island Health Authority (40/166, 24.1%). Also, an equal number were employed full-time (34/166, 20/5%) or unemployed (34/166, 20.5%) (Table 1).

**Table 1 –.** Demographic characteristics^1^.

| Category | Value | N | % |
| --- | --- | --- | --- |
| **Age** | 18-24 | 0/166 | 0% |
|  | 25 to 34 | 12/166 | 7.2% |
|  | 35 to 44 | 32/166 | 19.3% |
|  | 45 to 54 | 55/166 | 33.1% |
|  | 55 to 64 | 46/166 | 28% |
|  | 65 or older | 11/166 | 7% |
|  | Total respondents | 156/166 | 94% |
|  | No response | 10/166 | 6% |
| **Gender** | Woman | 112/166 | 67.5% |
|  | Man | 40/166 | 24.1% |
|  | Prefer not to answer | 2/166 | 1.2% |
|  | Transgender woman | 1/166 | 1% |
|  | Total respondents | 155/166 | 93.4% |
|  | No response | 11/166 | 7% |
| **Socioeconomic status** (total household income and other non-employment sources of income) | Very low-income | 9/166 | 5.4% |
|  | Low-income | 17/166 | 10.2% |
|  | Middle-income | 87/166 | 52.4% |
|  | High middle-income | 26/166 | 16% |
|  | High-income | 4/166 | 2.4% |
|  | Prefer not to answer | 12/166 | 7.2% |
|  | Total respondents | 155/166 | 93.4% |
|  | No Response | 11/166 | 7% |

<sup>1</sup> Some answers allow for multiple options, so totals do not always amount to 100%

|  |  |  |  |
| --- | --- | --- | --- |
| **Country of birth** | Canada | 123/166 | 74.1% |
|  | USA | 2/166 | 1.2% |
|  | Poland | 2/166 | 1.2% |
|  | UK | 6/166 | 4% |
|  | Other | 3/166 | 2% |
|  | Total respondents | 156/166 | 94% |
|  | No response | 10/166 | 6% |
| **Ethnic or Cultural Background** | Caucasian or White | 135/166 | 81.3% |
|  | Black | 1/166 | 1% |
|  | Indigenous | 6/166 | 4% |
|  | South Asian (Indian, Pakistani, Sri Lankan, etc.) | 4/166 | 2.4% |
|  | Japanese | 4/166 | 2.4% |
|  | Chinese | 4/166 | 2.4% |
|  | Filipino | 2/166 | 1.2% |
|  | Other | 8/166 | 5% |
|  | No response | 10/166 | 6% |
| **Domestic status** | Married or living with a partner | 107/166 | 65% |
|  | Single | 41/166 | 25% |
|  | Widow (er) | 3/166 | 2% |
|  | Other | 5/166 | 3% |
|  | Total respondents | 156/166 | 94% |
|  | No response | 10/166 | 6% |
| **Caretaking responsibilities** | Children / Stepchildren | 62/166 | 37.3% |
|  | Parents | 26/166 | 16% |
|  | None | 69/166 | 42% |
|  | Other | 12/166 | 7.2% |
|  | No response | 10/166 | 6% |
| **Education level** | Bachelor of Science in Nursing (BScN) or Bachelor of Nursing (BN) | 27/166 | 16.2% |
|  | Registered Nurse (RN) or Licensed Practical Nurse (LPN) or Registered Practical Nurse (RPN) diploma | 13/166 | 8% |
|  | Nursing Assistant Diploma | 9/166 | 5.4% |
|  | Master of Science in Nursing (MScN) | 1/166 | 1% |
|  | Doctor of Medicine (MD) | 11/166 | 7% |
|  | Paramedical or Emergency Medical Technician (EMT) program | 14/166 | 8.4% |
|  | Bachelor of Science (BSc) | 4/166 | 2.4% |
|  | Bachelor of Health Sciences (BHSc) | 2/166 | 1.2% |
|  | Master of Health Sciences (MHSc) | 4/166 | 2.4% |
|  | PhD (any field) | 1/166 | 1% |
|  | Bachelor of Health Science in Occupational Therapy | 1/166 | 1% |
|  | Master of Science in Occupational Therapy (MScOT) | 2/166 | 1.2% |
|  | Master of Social Worker (MSW) or Registered Social Worker (RSW) | 4/166 | 2.4% |
|  | Master of Health Sciences (MHSc) | 4/166 | 2.4% |
|  | Bachelor or advanced degree, Health Administration/ Systems Management (e.g., BHAD) | 2/166 | 1.2% |
|  | Bachelor or Doctor of Pharmacy | 2/166 | 1.2% |
|  | Registered Pharmacy Technician (RPhT) | 3/166 | 2% |
|  | Traditional Chinese Medicine (TCM) diploma | 2/166 | 1.2% |
|  | Acupuncture diploma | 1/166 | 1% |
|  | Phytotherapy (herbal medicine) diploma | 2/166 | 1.2% |
|  | Doctor of Naturopathy degree | 3/166 | 2% |
|  | Other | 70/166 | 42.2% |
|  | No response | 10/166 | 6% |
| **Profession/area of occupation** | Registered nurse/ Registered psychiatric nurse | 25/166 | 15.1% |
|  | Licensed practical nurse | 5/166 | 3% |
|  | Nurses aide/ Orderly/ Patient services associate (e.g., health care aide, long-term care aide, nursing assistant) | 16/166 | 10% |
|  | Nursing Coordinator/ Supervisor | 5/166 | 3% |
|  | Social Worker | 3/166 | 2% |
|  | Academic appointment/ University affiliation | 7/166 | 4.2% |
|  | In clinical practice | 30/166 | 18.1% |
|  | Not in clinical practice | 8/166 | 5% |
|  | Specialist physician | 6/166 | 4% |
|  | General practitioner/ Family physician | 4/166 | 2.4% |
|  | Allied primary health practitioner (e.g., nurse practitioner, midwife, physician assistant) | 1/166 | 1% |
|  | Paramedical occupation (e.g., EMT, ambulance attendant, advanced care paramedic) | 14/166 | 8.4% |
|  | Physiotherapist or Physiotherapy Assistant (PTA) | 1/166 | 1% |
|  | Occupational therapist or Occupational therapy Assistant (OTA) | 3/166 | 2% |
|  | Medical technologist/ Technician (e.g., medical laboratory technologist, respiratory therapist) | 9/166 | 5.4% |
|  | Pharmacist or Registered Pharmacy Technician | 4/166 | 2.4% |
|  | Dietician/ Nutritionist | 1/166 | 1% |
|  | Natural healing practitioner (e.g. acupuncturist, traditional Chinese medicine (TCM) practitioner) | 5/166 | 3% |
|  | Health Information Management | 2/166 | 1.2% |
|  | Other | 46/166 | 28% |
|  | No response | 10/166 | 6% |
| **Years of experience in most recent career** | 0-5 years | 19/166 | 11.4% |
|  | 6-10 years | 24/166 | 14.5% |
|  | 11-15 years | 31/166 | 19% |
|  | 16-20 years | 20/166 | 12% |
|  | 21-25 years | 23/166 | 14% |
|  | 26-30 years | 16/166 | 10% |
|  | 31-35 years | 15/166 | 9% |
|  | 36-39 years | 3/166 | 2% |
|  | 40+ years | 5/166 | 3% |
|  | Total respondents | 156/166 | 94% |
|  | No response | 10/166 | 6% |
| **Years of education/training** | 0-4 years | 50/166 | 30.1% |
|  | 5-9 years | 38/166 | 23% |
|  | 10+ years | 68/166 | 41% |
|  | Total respondents | 156/166 | 94% |
|  | No response | 10/166 | 6% |
| **British Columbia health affiliation<sup>2</sup>** | Fraser Health | 40/166 | 24.1% |
|  | Interior Health | 49/166 | 29.5% |
|  | Northern Health | 13/166 | 8% |
|  | Vancouver Coastal Health | 15/166 | 9% |
|  | Vancouver Island Health Authority | 40/166 | 24.1% |
|  | Provincial Health Services Authority (PHSA) | 11/166 | 7% |
|  | Other | 11/166 | 7% |
|  | No response | 10/166 | 6% |
| **Employment status** | Employed full-time | 34/166 | 20.5% |
|  | Employed part-time | 30/166 | 18.1% |
|  | Unemployed | 34/166 | 20.5% |
|  | Retired | 10/166 | 6% |
|  | Self-employed | 28/166 | 17% |
|  | Casual | 4/166 | 2.4% |
|  | Contractor | 4/166 | 2.4% |
|  | Other | 12/166 | 7.2% |
|  | Total respondents | 156/166 | 94% |
|  | No response | 20/166 | 6% |

<sup>2</sup> Government of British Columbia. (no date). *Health Authorities - Province of British Columbia*. Retrieved Septemebr 9, 2024, from https://www2.gov.bc.ca/gov/content/health/about-bc-s-health-care-system/partners/health-authorities

### Vaccination decision and experiences

Most respondents (143/166, 86.1%) were not vaccinated (Figure 1). Of those who were vaccinated, more than half had received a partial primary series (7/13, 54%), over one third a complete primary series (5/13, 38.5%), a small minority (1/13, 8%) had been boosted once, and no one reported being boosted twice or more times (0/13, 0%). Most (10/13, 78%) were vaccinated primarily because it was mandated for work, and a small minority (2/13, 15.4%) to protect the larger community (1/13, 8%) or themselves (1/13, 8%) from severe outcomes. Most vaccinated respondents (10/13, 78%) did not recommend the vaccine to others and most (11/13, 85%) reported experiencing adverse effects post vaccination, whereas a small number (2/13, 15.4%) reported no such events (National Cancer Institute, 2021). Adverse effects were mild after the 1^st^ (3/13, 23.1%) and the 2^nd^ doses (2/13, 15.4%); moderate after 1^st^ (3/13, 23.1%) and the 2^nd^ doses (1/13, 8%), or severe after the 1^st^ (3/13, 23.1%) and the 2^nd^ doses (1/13, 8%). About one-third of vaccinated respondents (4/13, 31%) did not communicate their reaction to a doctor, while almost half (6/13, 46.2%) did. Among these, in only a minority of the cases (1/6, 17%) a report was filed, in half of them no report was filed (3/6, 50%), and in one-third of them (2/6, 33.3%) respondents did not know if a report had been filed. More than half (7/13, 54%) of vaccinated respondents reported that after experiencing an adverse reaction their employer had still required additional doses. Only a small number (2/13, 15.4%) reported no adverse events post vaccination (Table 2).

**Chart 1.**
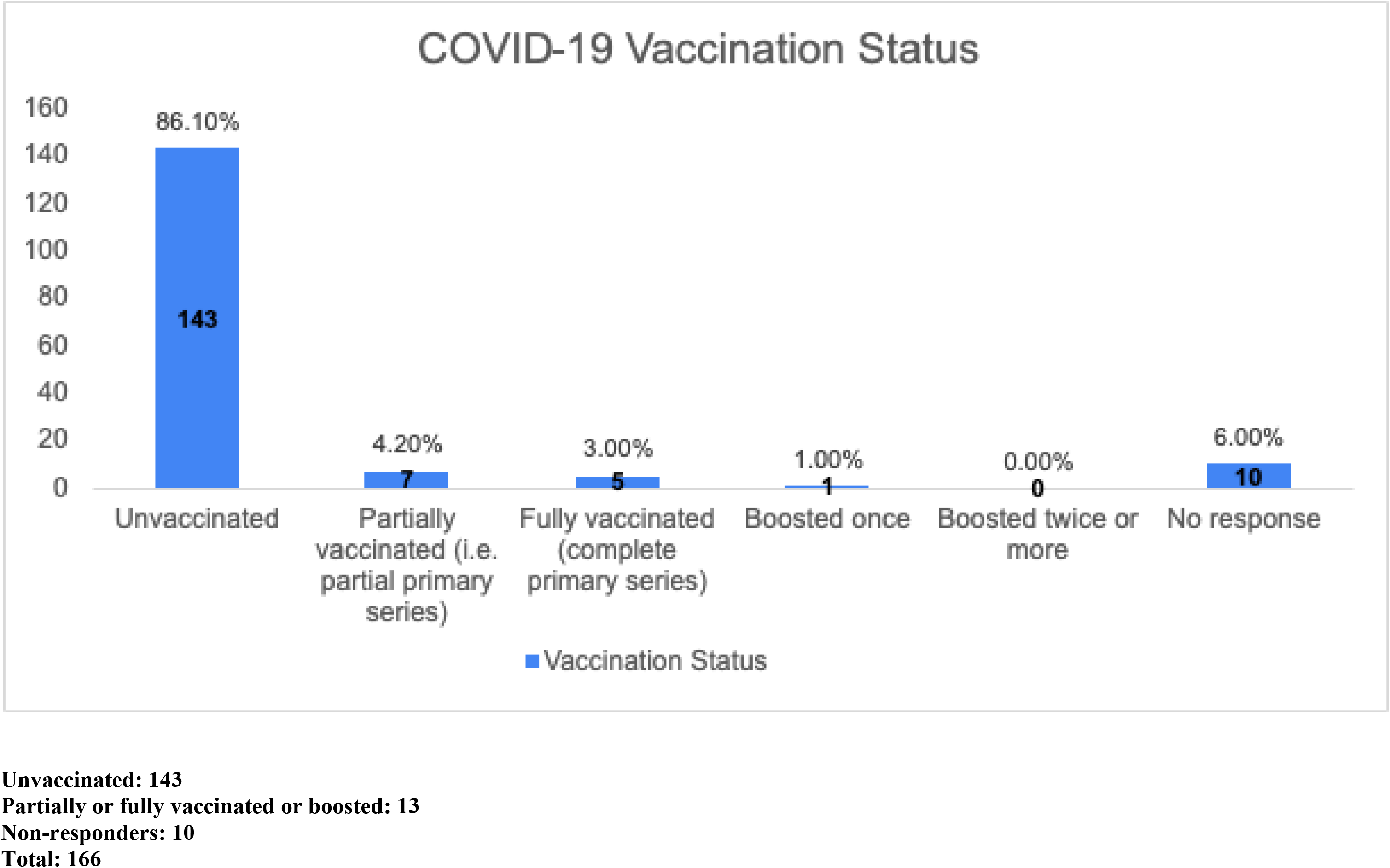
Vaccination status.

**Table 2 –.**
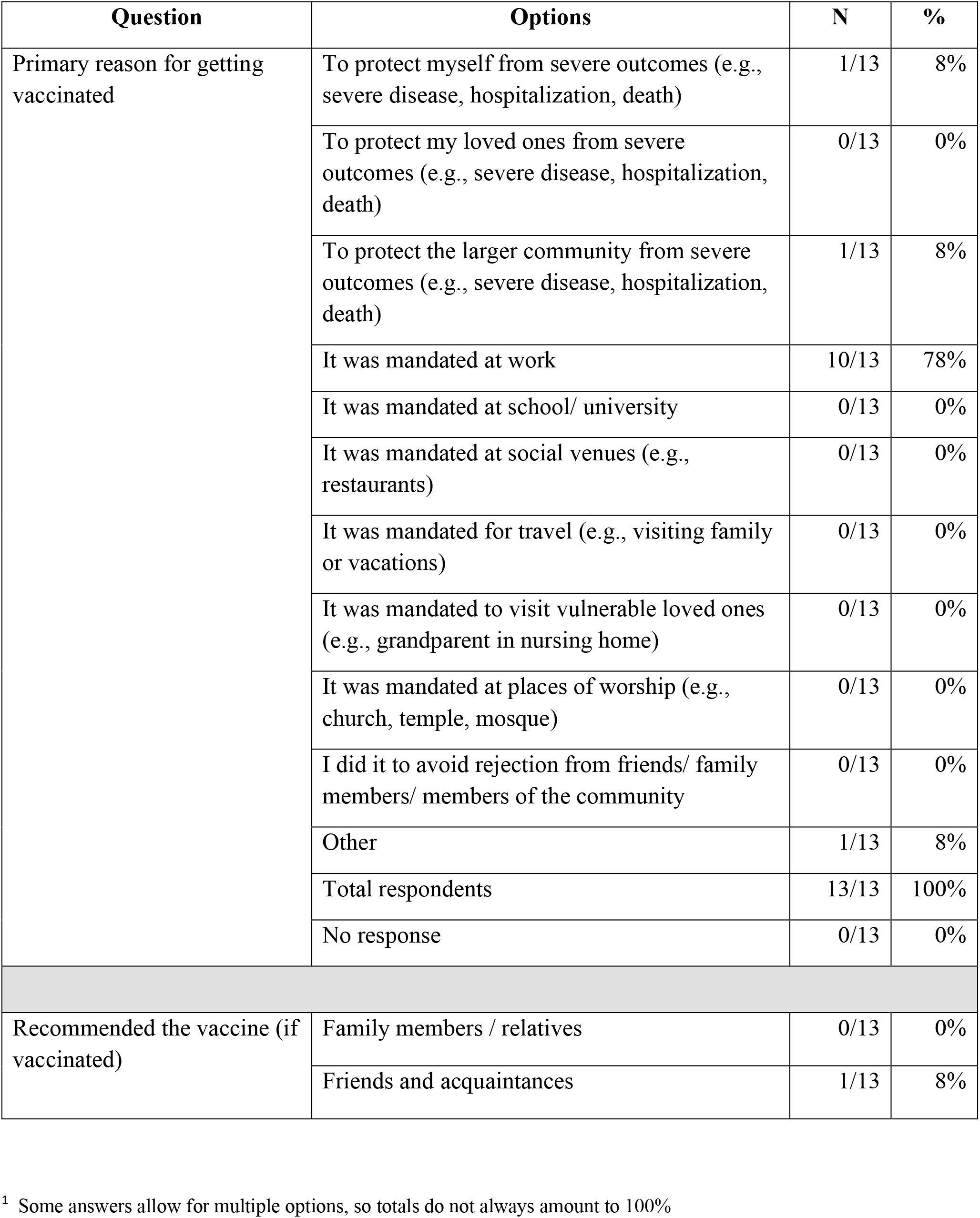
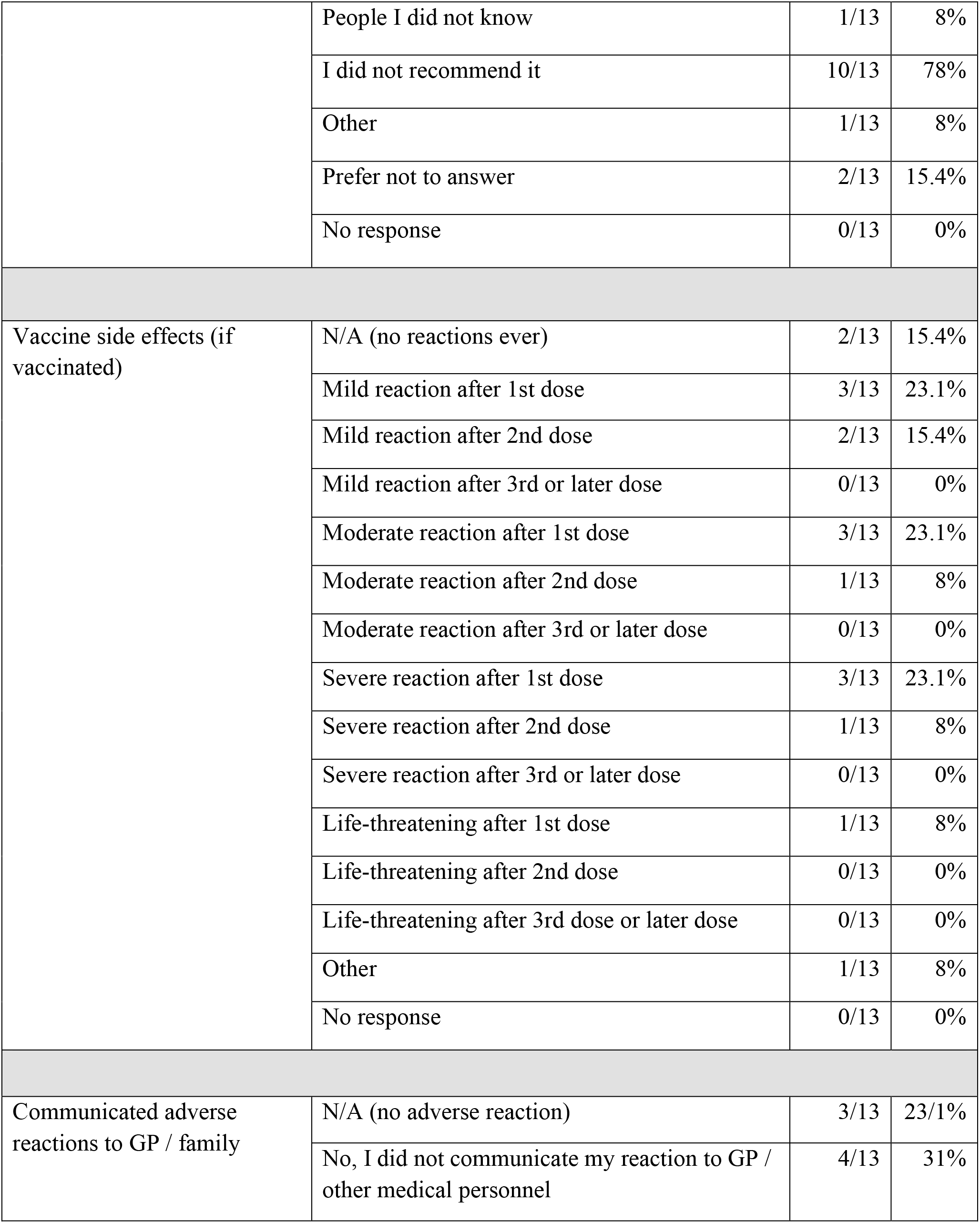
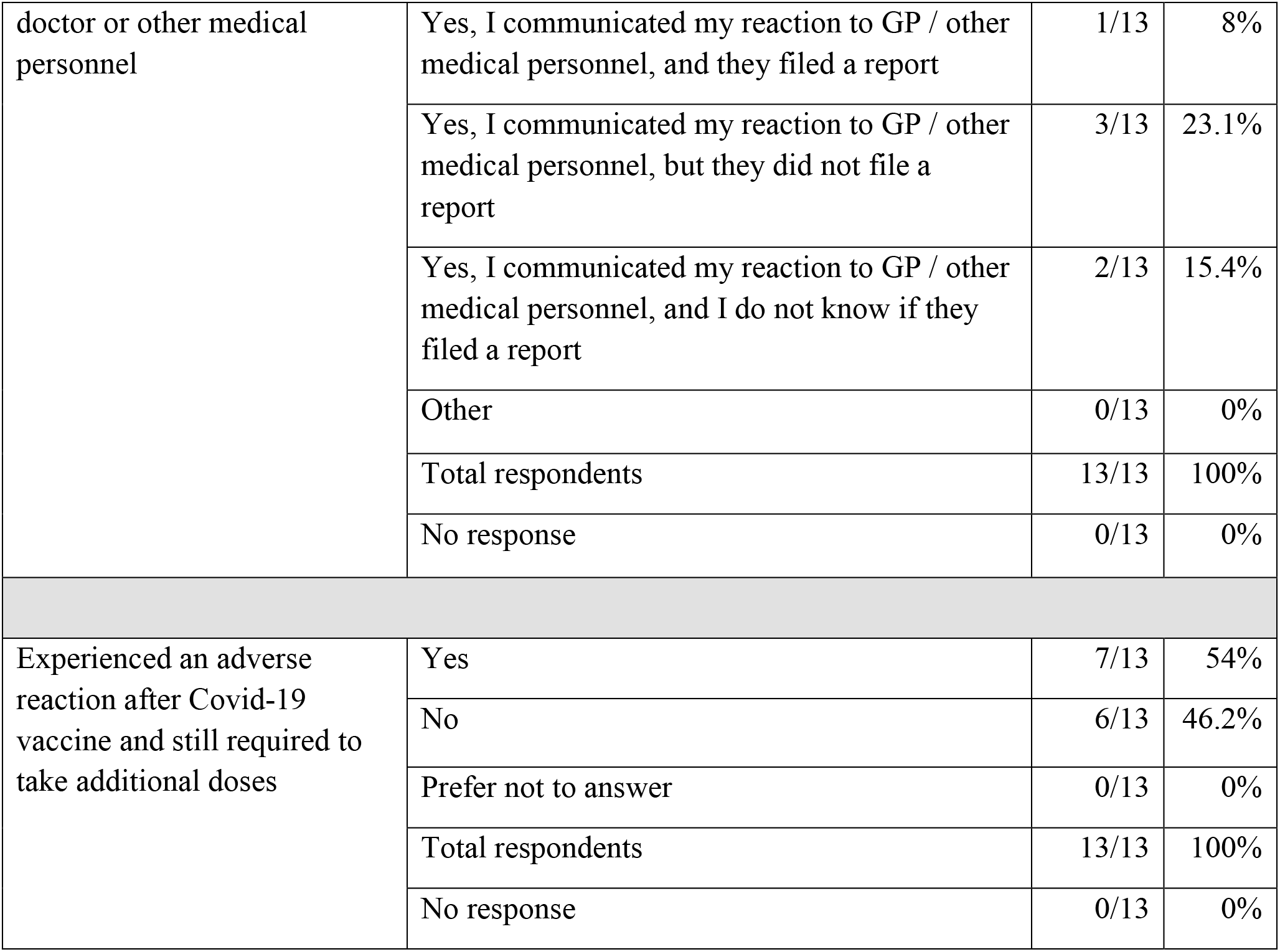
Vaccination decision and experience^1^.

### Personal and family impact of vaccination policies

Most respondents (121/166, 73%) agreed (17/166, 10.2%) or strongly agreed (104/166, 63%) that their current income was lower than it was prior to the introduction of vaccination mandates. Not surprisingly, most laid off respondents (120/138, 87%) also agreed (12/138, 9%) or strongly agreed (108/138, 78.3%) that being terminated had significantly reduced their income. Responses to the impact of vaccination policies on physical health were more mixed. About half (81/166, 49%) of all respondents chose “not applicable”— unsurprisingly since most respondents were unvaccinated. A minority (33/166, 20%), however, reported that they agreed (11/166, 7%) or strongly agreed (22/166, 13.3%) that their physical health had worsened after mandates were implemented, while the remainder (40/166, 24.1%) were neutral (8/166, 5%), disagreed (18/166, 11%) or strongly disagreed (14/166, 8.4%) (Table 3). Similarly, answers to whether respondents had suffered physical disabilities due to vaccination requirements were also mixed, with close to one-fourth (40/166, 24.1%) disagreeing (20/166, 12%) or strongly disagreeing (20/166, 12%) that they had, about one-tenth (15/166, 9%) agreeing (5/166, 3%) or strongly agreeing (10/166, 6%), and a slightly smaller percentage (9/166, 5.4%) remaining neutral. In contrast, most respondents (135/166, 81.3%) reported experiencing anxiety or depression due to mandates, with close to one-fourth (39/166, 23.5%) agreeing (15/166, 9%) or strongly agreeing (24/166, 14.5%) that they had experienced suicidal thoughts due to employer vaccination requirements, and close to half (76/166, 46%) agreeing (22/166, 13.3%) or strongly agreeing (54/166, 32.5%) that they had sought help from a counsellor due to situations arising from these requirements.

**Table 3 -.** Personal and family impact of vaccination policies^1^.

| **Statement** | **Strongly disagree**<br>**(N; %)** | **Disagree**<br>**(N; %)** | **Neutral**<br>**(N; %)** | **Agree**<br>**(N; %)** | **Strongly agree**<br>**(N; %)** | **N. A** | **No response** |
| --- | --- | --- | --- | --- | --- | --- | --- |
| My income is less than it was prior to the introduction of vaccination policies / mandates | 14/166;<br>8.4% | 11/166;<br>7% | 5/166;<br>3% | 17/166;<br>10.2% | 104/166;<br>63% | 4/166;<br>2.4% | 11/166;<br>7% |
| Losing my job significantly reduced my income | 4/138;<br>3% | 6/138;<br>4.3% | 5/138;<br>4% | 12/138;<br>9% | 108/138;<br>78.3% | 3/138;<br>2.2% | 0/138;<br>0% |
| I have suffered chronic physical ailments due to employer vaccination requirements | 14/166;<br>8.4% | 18/166;<br>11% | 8/166;<br>5% | 11/166;<br>7% | 22/166;<br>13.3% | 81/166;<br>49% | 12/166;<br>7.2% |
| I have suffered physical disability due to employer vaccination requirements | 20/166;<br>12% | 20/166;<br>12% | 9/166;<br>5.4% | 5/166;<br>3% | 10/166;<br>6% | 90/166;<br>54.2% | 12/166;<br>7.2% |
| I have suffered anxiety and/ or depression due to employer vaccination requirements | 7/166;<br>4.2% | 2/166;<br>1.2% | 5/166;<br>3% | 30/166;<br>18.1% | 105/166;<br>63.3% | 6/166;<br>4% | 11/166;<br>7% |
| I have experienced suicidal thoughts due to employer vaccination requirements | 39/166;<br>23.5% | 28/166;<br>17% | 20/166;<br>12% | 15/166;<br>9% | 24/166;<br>14.5% | 29/166;<br>17.5% | 11/166;<br>7% |
| I have sought help from a counsellor due to situations arising from vaccination requirements | 26/166;<br>16% | 18/166;<br>11% | 10/166;<br>6% | 22/166;<br>13.3% | 54/166;<br>32.5% | 25/166;<br>15.1% | 11/166;<br>7% |
| My personal relationships (spouses, friends) suffered due to situations arising from vaccination requirements | 4/166;<br>2.4% | 4/166;<br>2.4% | 6/166;<br>4% | 26/166;<br>16% | 113/166;<br>68.1% | 2/166;<br>1.2% | 11/166;<br>7% |

<sup>1</sup> The denominator of 138 is comprised of respondents who replied “yes/prefer not to answer” when asked whether they were terminated as shown in Table 4

|  |  |  |  |  |  |  |  |
| --- | --- | --- | --- | --- | --- | --- | --- |
| I feel I have been unfairly treated by my employer regarding vaccination requirements. | 2/166;<br>1.2% | 5/166;<br>3% | 2/166;<br>1.2% | 8/166;<br>5% | 137/166;<br>82.5% | 0/166;<br>0% | 12/166;<br>7.2% |
| Losing my job had a negative impact on my physical health | 4/138;<br>3% | 17/138;<br>12.3% | 19/138;<br>14% | 35/138;<br>25.4% | 59/138;<br>43% | 4/138;<br>3% | 0/138;<br>0% |
| Losing my job had a negative impact on my mental health | 0/138;<br>0% | 5/138;<br>4% | 11/138;<br>8% | 31/138;<br>22.5% | 88/138;<br>64% | 3/138;<br>2.2% | 0/138;<br>0% |

As well, most respondents (139/166, 84%) agreed (26/166, 16%) or strongly agreed (113/166, 68.1%) that their personal relationships had suffered due to situations arising from mandated vaccination, with most (119/138, 86.2%) non-compliant respondents agreeing (31/138, 22.5%) or strongly agreeing (88/138, 64%) that being terminated had a negative impact on their mental health. Most respondents (145/166, 87.3%) also agreed (8/166, 5%) or strongly agreed (137/166, 82.5%) with the statement “I feel I have been unfairly treated by my employer regarding vaccination requirements” (Table 3). Finally, while about half of respondents (85/166, 51.2%) reported good (32/166, 19.3%) or very good (53/166, 32%) physical health, over one-third (57/166, 34.3%) reported experiencing better physical health before COVID-19. This change was even more marked for mental health, that most (86/166, 52%) respondents rated as good (48/166, 29%) or very good (38/166, 23%), yet most (97/166, 58.4%) also rated their mental health as having been better before COVID-19 (Figures 2,3).

**Chart 2 –.**
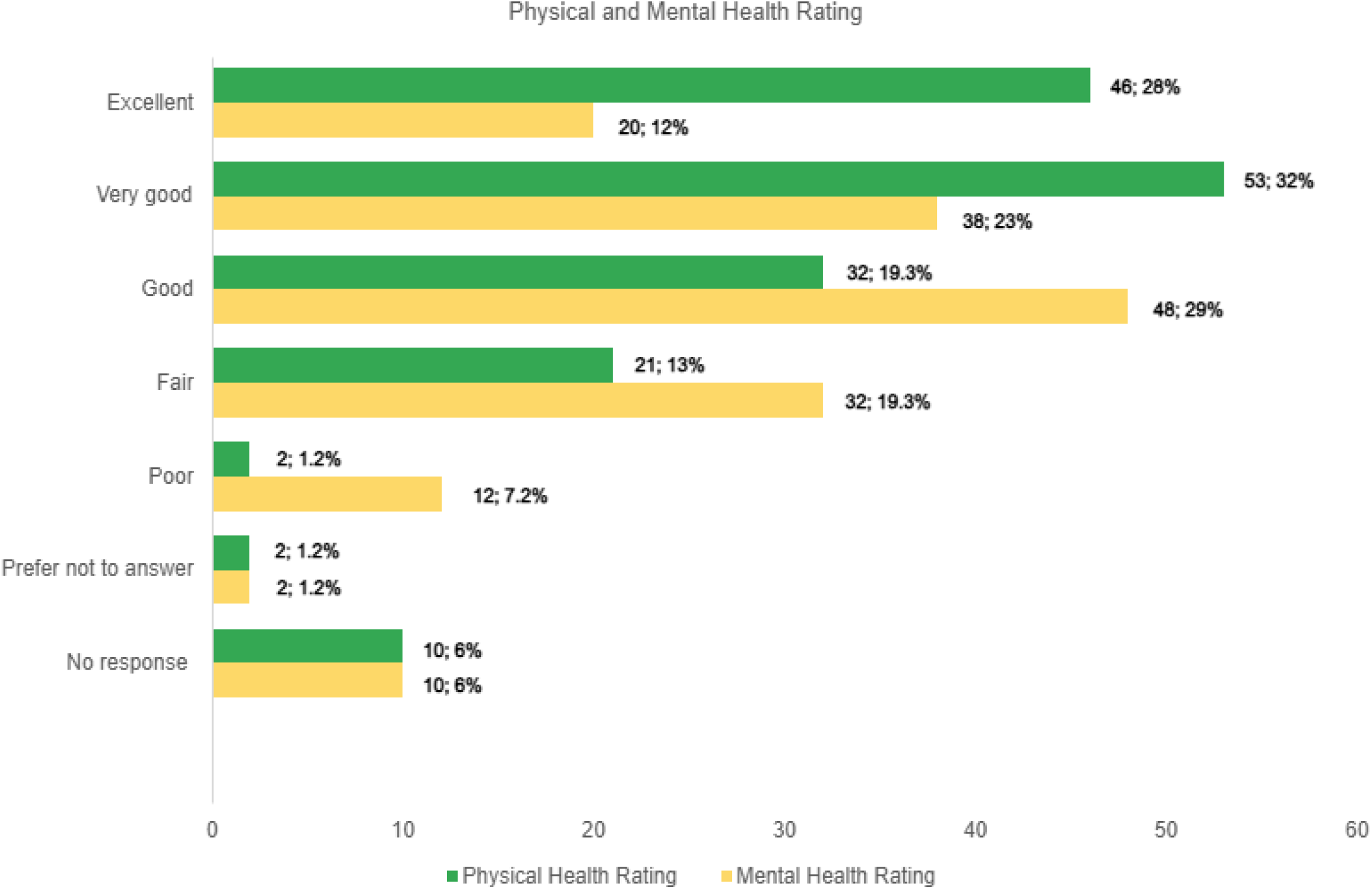
Physical and mental health self-rating.

**Chart 3 –.**
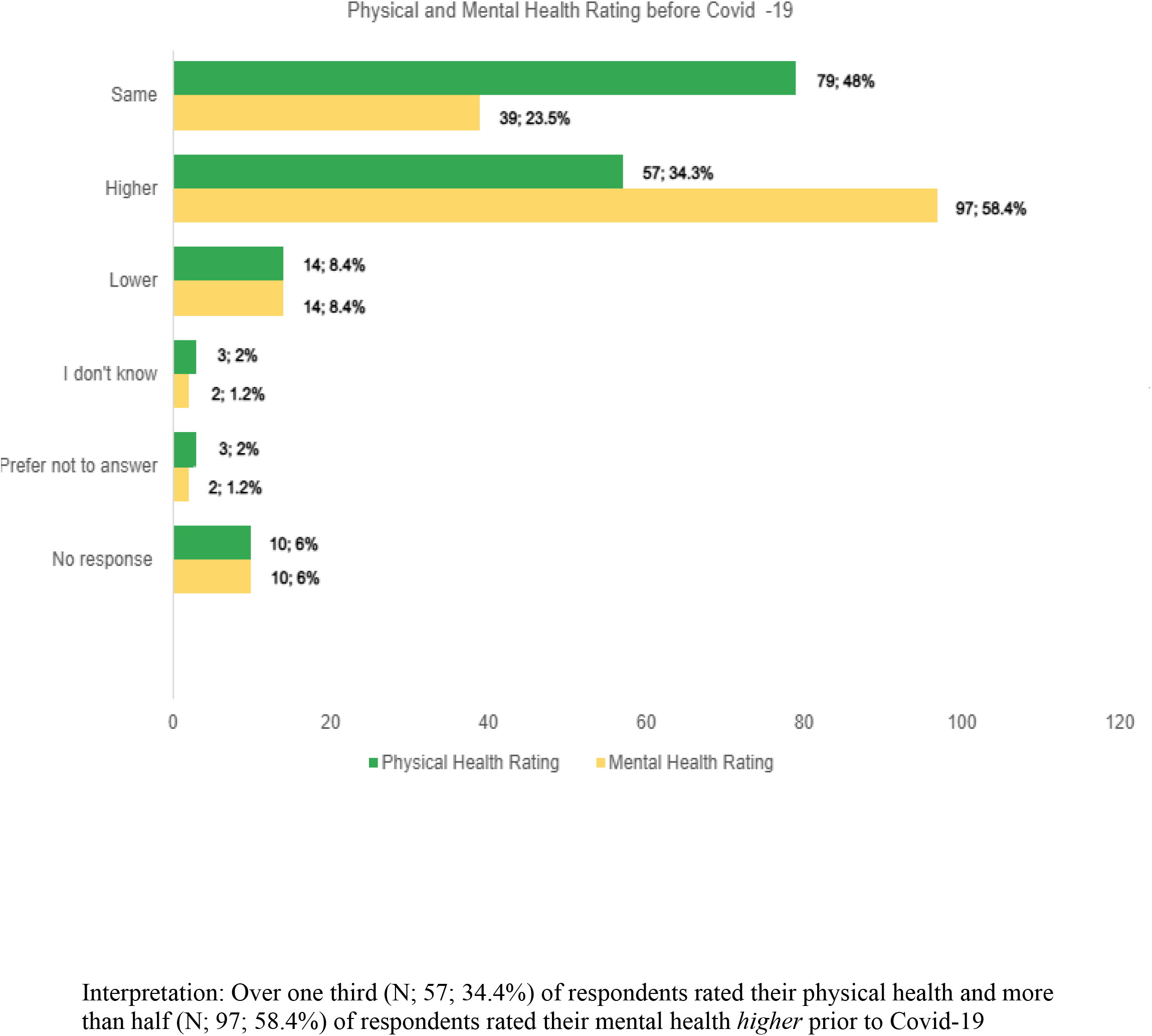
Physical and mental health self-rating before Covid-19.

### Workplace and labour market impact of vaccination policies

Nearly three-quarters (133/166, 80.1%) of respondents reported that they had been terminated due to their decision to not be vaccinated, either not at all or after one or two doses (i.e., booster mandates). In addition, close to half (70/166, 42.2%) reported that they had been subjected to disciplinary measures other than termination, such as accusations of professional misconduct, reports to licensing colleges, temporary suspension of pay, exclusion from pension plans, or withdrawal of their professional license (Table 4). Finally, most respondents (132/166, 79.5%) agreed (25/166, 17%) or strongly agreed (107/166, 64.5%) that after the introduction of vaccines or vaccination policies they had experienced conflict among colleagues. Most (131/166, 79%) also agreed (23/166, 14%) or strongly agreed (108/166, 65.1%) that after vaccine mandates were introduced, they had experienced conflict between employees and management. As well, most respondents (139/166, 84%) reported knowing of HCWs who had taken early retirement due to COVID-19 policies, most (141/166, 85%) knew of colleagues who had been laid off due to non-compliance with vaccine mandates, and most (138/166, 83.1%) knew of colleagues who had resigned because they did not wish to take the vaccine (Table 5).

**Table 4 –.** Vaccination requirements & impact on employment status and conditions.

| Question | Options | N | % |
| --- | --- | --- | --- |
| Terminated or laid off due to the decision to not receive the Covid-19 vaccine (first or subsequent doses)? | Yes | 133/166 | 80.1% |
|  | No | 17/166 | 10.2% |
|  | Prefer not to answer | 5/166 | 3% |
|  | Total respondents | 155/166 | 93.4% |
|  | No response | 11/166 | 7% |
| Subject to disciplinary measures *other* than layoffs (e.g., accusations of “professional misconduct”; reports to licensing colleges; temporary suspension of pay; exclusion from pension plan; withdrawal of professional license). | Yes | 70/166 | 42.2% |
|  | No | 62/166 | 37.3% |
|  | Prefer not to answer | 8/166 | 5% |
|  | Other | 15/166 | 9% |
|  | No response | 11/166 | 7% |

**Table 5 –.**
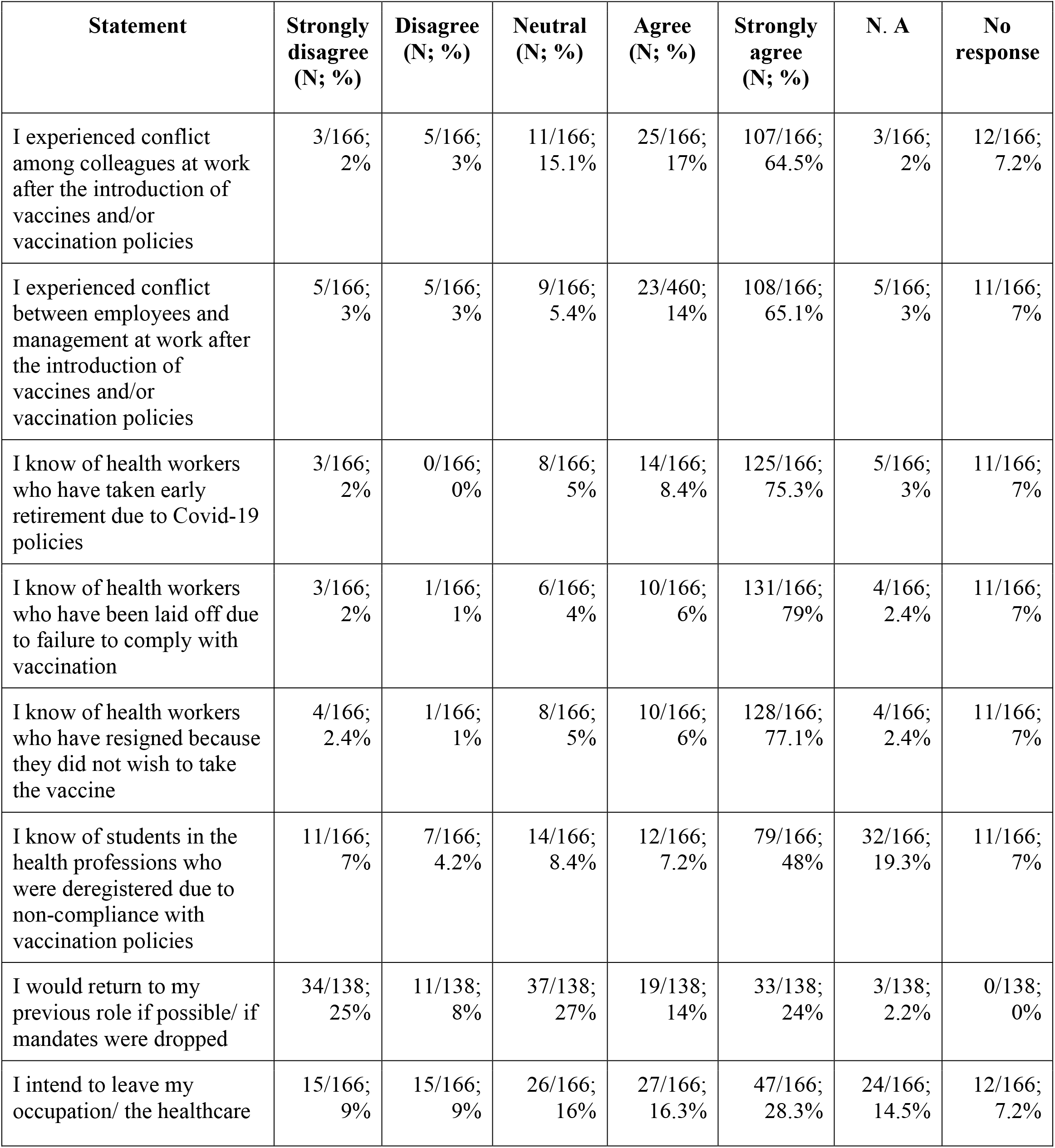
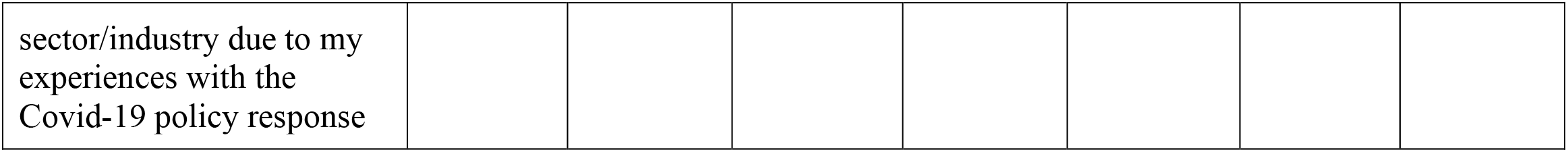
Level of agreement with statements on vaccination requirements & impact on employment status and conditions.

In addition, more than half (91/166, 55%) of respondents knew of students in the health professions who were de-enrolled by their educational institutions due to non-compliance with vaccination policies. The response to the statement “I would return to my previous role if possible/if mandates were dropped” was split. While a large minority (52/138, 38%) agreed (19/138, 14%) or strongly agreed (33/138, 24%) that they would, about the same proportion (45/138, 33%) disagreed (11/138, 8%) or strongly disagreed (34/138, 25%), while a small minority (37/138, 27%) was neutral. Finally, respondents reported mixed feelings about remaining employed in healthcare: nearly half (74/166, 44.6%) agreed (27/166, 16.3%) or strongly agreed (47/166, 28.3%) that they intended to leave their occupation or the healthcare sector altogether due to their experiences with Covid-19 policies, while close to one-fifth (30/166, 18.1%) reported no plans to leave the industry, and about the same proportion (26/166, 19%) were neutral (Table 5).

### Accommodation, equity considerations & informed consent

Most respondents (155/166, 93.4%) reported that they were not offered any alternatives to vaccination. Nearly half (73/166, 44%) requested, but did not receive, an exemption. The most common reason for requesting an exemption was religious (41/166, 25%), followed by medical (38/166, 23%) and conscientious objection (16/166, 10%) grounds. However, over one-fourth of respondents (45/166, 27.1%) reported that they did not request an exemption because they were not eligible or felt intimidated or discouraged by the rejection of other HCWs’ requests. When asked if their employer (or professional college or public health authority if self-employed) had provided them with written information about COVID-19 vaccines, most respondents (119/166, 72%) reported that they had not, about one-sixth (27/166, 16.3%) reported that they were provided information from public health agencies or equivalent, and no respondents (0/166, 0%) reported being provided a package insert from the vaccine manufacturer. About one-fifth (33/166, 20%) reported that, if received, the information from employers had not enabled them to make an informed decision about vaccination (Table 6).

**Table 6 –.** Accommodations, EDI considerations & informed consent.

| **Question** | **Options** | **N** | **%** |
| --- | --- | --- | --- |
| Employer or regulatory authorities offered alternatives to vaccination | Yes, testing on site paid for by employer | 1/166 | 8% |
|  | Yes, test off site at your cost | 0/166 | 0% |
|  | Yes, remote work | 0/166 | 0% |
|  | Yes, educational training | 0/166 | 0% |
|  | Yes, proof of natural immunity | 0/166 | 0% |
|  | No, they did not offer any alternatives to vaccination | 155/166 | 93.4% |
|  | Total respondents | 156/166 | 94% |
|  | No response | 10/166 | 6% |
| Requested exemption from vaccination | N/A (e.g., my employer did not request mandatory vaccination, and I did not need an exemption) | 3/166 | 2% |
|  | No, I was not interested in requesting an exemption | 10/166 | 6% |
|  | Yes, and I received an exemption | 1/166 | 1% |
|  | Yes, but I did not receive an exemption | 73/166 | 44% |
|  | No, I did not request an exemption (e.g., because I was intimidated to ask for one) | 45/166 | 27.1% |
|  | Other | 9/166 | 14.5% |
|  | No response | 10/166 | 6% |
| Category of exemption | N/A (did not apply for exemption) | 74/166 | 44.6% |
|  | Medical | 38/166 | 23% |
|  | Religious | 41/166 | 25% |
|  | Conscientious | 16/166 | 10% |
|  | Other | 26/166 | 16% |
|  | No response | 12/166 | 7.2% |
| Employer (or professional college or public health authority if self-employed) provided written information about the vaccines | Yes, I was provided a package insert from the vaccine manufacturer(s) | 0/166 | 0% |
|  | Yes, I was provided information from public health agencies or equivalent | 27/166 | 16.3% |
|  | No, they never provided me with written information about the vaccines | 119/166 | 72% |
|  | Other | 10/166 | 6% |
|  | No response | 11/166 | 7% |
| If you received written information from your employer (or professional college or public health authority if self-employed) did it enable you to make an informed decision about vaccination? | Yes | 9/166 | 5.4% |
|  | No | 33/166 | 20% |
|  | I did not receive written information | 99/166 | 60% |
|  | Other | 14/166 | 8.4% |
|  | Total respondents | 155/166 | 93.4% |
|  | No response | 11/166 | 7% |

When asked their level of agreement with various statements related to informed consent, most respondents (149/166, 90%) disagreed (9/166, 5.4%) or strongly disagreed (140/166, 84.3%) that they had felt fully free to get or not get vaccinated, and most (119/166, 72%) also disagreed (15/166, 9%) or strongly disagreed (104/166, 63%) that they felt comfortable sharing their concerns about vaccination with their employer. In contrast, most (151/166, 91%) agreed (1/166, 1%) or strongly agreed (150/166, 90.4%) that they had safety concerns with Covid-19 vaccines, and more than half (96/166, 58%) agreed (5/166, 6%) or strongly agreed (91/166, 45.4%) that they had medical concerns, while more than half (95/166, 57.2%) agreed (16/166,10%) or strongly agreed (79/166, 47.6%) that they had religious concerns regarding the Covid-19 vaccines. Most (139/166, 84%) also agreed (13/166, 18%) or strongly agreed (126/166, 76%) that they did their own research regarding the safety and efficacy of Covid-19 vaccines. As well, close to one-fifth of respondents of all vaccination statuses (51/166, 31%) strongly agreed that they had felt coerced to get vaccinated. Finally, most vaccinated respondents (11/13, 85%) disagreed (1/13, 8%) or strongly disagreed (10/13, 77%) that they were happy about getting vaccinated, whereas most unvaccinated respondents (134/143, 94%) agreed (2/143, 1.4%) or strongly agreed (132/143, 92.3%) that they were happy to have remained unvaccinated (Table 7).

**Table 7 –.** Level of agreement with statements on vaccine concerns & informed consent.

| **Statement**<br>Level of agreement with the following related to the decision on Covid-19 vaccines | **Strongly disagree**<br>(N; %) | **Disagree**<br>(N; %) | **Neutral**<br>(N; %) | **Agree**<br>(N; %) | **Strongly agree**<br>(N; %) | **N. A**<br>(N; %) | **No response** |
| --- | --- | --- | --- | --- | --- | --- | --- |
| I felt entirely free to choose whether or not to get vaccinated | 140/166;<br>84.3% | 9/166;<br>5.4% | 3/166;<br>2% | 2/166;<br>1.2% | 1/166;<br>1% | 0/166;<br>0% | 11/166<br>; 7% |
| I had safety concerns with the Covid-19 vaccines | 4/166;<br>2/4% | 0/166;<br>0% | 0/166;<br>0% | 1/166;<br>1% | 150/166;<br>90.4% | 0/166;<br>0% | 11/166<br>; 7% |
| I had personal medical concerns with the Covid-19 vaccines (e.g., I have an autoimmune disorder) | 8/166;<br>9% | 6/166;<br>6.2% | 11/166;<br>9.2% | 5/166;<br>6% | 91/166;<br>45.4% | 33/166;<br>20% | 12/166<br>; 7.2% |
| I had religious concerns with the Covid-19 vaccines. | 17/166;<br>10.2% | 16/166;<br>10% | 11/166;<br>7% | 16/166;<br>10% | 79/166;<br>47.6% | 25/166;<br>15.1% | 12/166<br>; 7.2% |
| I felt comfortable expressing safety concerns about the Covid-19 vaccines with my employer | 104/166;<br>63% | 15/166;<br>9% | 2/166;<br>1.2% | 6/166;<br>4% | 23/166;<br>14% | 4/166;<br>2.4% | 12/166<br>; 7.2% |
| I did my own research to determine the safety and efficacy of the Covid-19 vaccines | 3/166;<br>2% | 2/166;<br>1.2% | 7/166;<br>4.2% | 13/166;<br>18% | 126/166;<br>76% | 3/166;<br>2% | 12/166<br>7.2% |
| I felt coerced to get vaccinated | 2/166;<br>12% | 1/166;<br>1% | 1/166;<br>1% | 0/166;<br>0% | 51/166;<br>31% | 99/166;<br>60% | 12/166<br>; 7.2% |
| I am happy with my choice to get vaccinated (if you did not get vaccinated choose N/A) | 10/13;<br>77% | 1/13;<br>8% | 0/13;<br>0% | 0/13;<br>0% | 2/13;<br>15.4% | 141/143;<br>99% | 12/166<br>; 7.2% |
| I am happy with my choice to NOT get vaccinated (if you got vaccinated choose NA) | 2/143;<br>1.4% | 0/143;<br>0% | 2/143;<br>1.4% | 2/143;<br>1.4% | 132/143;<br>92.3% | 13/13;<br>1% | 15/166<br>; 9% |

### HCWs views and experiences of mandates on patient care

More than half of respondents (95/166, 57.2%) had worked with Covid-19 positive or suspected patients prior to the vaccine mandate (Table 8), and most (112/166, 67.5%) agreed (26/166, 16%) or strongly agreed (86/166, 52%) that they had observed concerning patient care or procedural changes upon the onset of Covid-19 (Table 9). Similarly, most (121/166, 73%) agreed (27/166, 16.3%) or strongly agreed (94/166, 57%) that they had observed disturbing patient care or procedural changes upon the introduction of Covid-19 vaccines, most (118/166, 71.1%) agreed (22/166, 13.3%) or strongly agreed (96/166, 58%) that they had observed differential treatment of patients based on their vaccination status, and most (117/166, 70.5%) agreed (25/166, 15.1%) or strongly agreed (92/166, 55/4%) that they had observed an increase in patient harms associated with the Covid-19 vaccine. Significantly, only a very small minority (8/166, 5%) agreed (1/166, 1%) or strongly agreed (7/166, 4.2%) that they had felt free to express to their employer their concerns about potential vaccine harms in patients, only a very small minority (11/166, 7%) agreed (4/166, 2.4%) or strongly agreed (7/166, 4.2%) that when they had expressed these concerns they had been documented or acted upon by their employer, and only a small minority (8/166, 5%) agreed (1/166, 1%) or strongly agreed (7/166; 4.2%) that from the perspective of a potential patient, they felt confident that the healthcare system would provide adequate and quality care while respecting their personal preferences and values (Table 9).

**Table 8 –.** Impact on patient care.

| **Question** | **Options** | **N** | **%** |
| --- | --- | --- | --- |
| Worked with Covid-19 positive or suspected patients' pre-vaccine mandate | Yes | 95/166 | 57.2% |
|  | No | 26/166 | 16% |
|  | Not sure | 25/166 | 15.1% |
|  | Other | 10/166 | 6% |
|  | Total respondents | 156/166 | 94% |
|  | No response | 10/166 | 6% |
| Encouraged to report adverse events post vaccination if observed | Yes | 3/166 | 2% |
|  | No | 152/166 | 92% |
|  | Total respondents | 155/166 | 94% |
|  | No response | 11/166 | 7% |
| Trained to report adverse events post-vaccination if observed | Yes | 16/166 | 10% |
|  | No | 138/166 | 83.1% |
|  | Total | 154/166 | 93% |
|  | No response | 12/166 | 7.2% |
| Asked, encouraged, or coerced to minimize vaccine hesitancy by: | Not providing exemptions when requested by patients | 12/166 | 7.2% |
|  | Not prescribing off label prescriptions | 13/166 | 8% |
|  | Telling patients that the vaccines were safe and effective | 44/166 | 27% |
|  | Encouraging patients to trust health officials and sources | 51/166 | 31% |
|  | Dismissing non-officially approved information as 'misinformation' | 38/166 | 23% |
|  | I was not asked to do any of these things | 65/166 | 39.1% |
|  | Other | 14/166 | 8.4% |
|  | Prefer not to answer | 18/166 | 11% |
|  | Total respondents | 154/166 | 93% |
|  | No response | 12/166 | 7.2% |
| Personally administered Covid-19 vaccines | Yes | 2/166 | 1.2% |
|  | No | 153/166 | 92.2% |
|  | Total respondents | 155/166 | 94% |
|  | No response | 11/166 | 7% |
| If administered Covid-19 vaccines, received compensation | Yes | 1/2 | 50% |
|  | No | 1/2 | 50% |
|  | Total respondents | 2/2 | 100% |
| Feeling upon administering Covid-19 vaccines | Accepting; it was part of my responsibility | 0/2 | 0% |
|  | Energized; I was part of the solution for a serious public health problem | 0/2 | 0% |
|  | Uneasy; I did not know what might happen to vaccine recipients | 2/2 | 100% |
|  | Other | 0/2 | 0% |
|  | Prefer not to answer | 0/2 | 0% |
|  | Total respondents | 2/2 | 100% |
|  | No response | 0/2 | 0% |

**Table 9 –.** Level of agreement with statements on impact on patient care.

| **Statement** | **Strongly disagree (N; %)** | **Disagree (N; %)** | **Neutral (N; %)** | **Agree (N; %)** | **Strongly agree (N; %)** | **N. A** | **No response** |
| --- | --- | --- | --- | --- | --- | --- | --- |
| I observed concerning patient care or procedural changes coinciding with the onset of the Covid-19 crises | 5/166;<br>3% | 4/166;<br>2.4% | 19/166;<br>11.4% | 26/166;<br>16% | 86/166;<br>52% | 15/166;<br>9% | 11/166;<br>7% |
| I observed concerning patient care changes after the introduction of the Covid-19 vaccines | 3/166;<br>2% | 4/166;<br>2.4% | 11/166;<br>7% | 27/166;<br>16.3% | 94/166;<br>57% | 16/166;<br>10% | 11/166;<br>7% |
| I observed differential treatment of patients based on their vaccine status | 2/166;<br>1.2% | 7/166;<br>4.2% | 14/166;<br>8.4% | 22/166;<br>13.3% | 96/166;<br>58% | 14/166;<br>8.4% | 11/166;<br>7% |
| I observed an increase in patient harms associated with the Covid-19 vaccines | 1/166;<br>1% | 4/166;<br>2.4% | 17/166;<br>10.2% | 25/166;<br>15.1% | 92/166;<br>55.4% | 15/166;<br>9% | 12/166;<br>7.2% |
| I felt free to express any concerns I had about patient care or potential vaccine harms with my employer | 107/166;<br>64.5% | 19/166;<br>11.4% | 8/166; 5% | 1/166;<br>1% | 7/166;<br>4.2% | 13/166;<br>8% | 11/166;<br>7% |
| If I expressed concerns about patient care or potential vaccine harms, these concerns were documented and acted upon by my employer | 58/166;<br>35% | 25/166;<br>15.1% | 23/166;<br>14% | 4/166;<br>2.4% | 7/166;<br>4.2% | 38/166;<br>23% | 11/166;<br>7% |
| From the perspective of a potential patient, I am confident that the current healthcare system will provide adequate and | 118/166;<br>71.1% | 19/166;<br>11.4% | 9/166;<br>5.4% | 1/166;<br>1% | 7/166;<br>4.2% | 2/166;<br>1.2% | 11/166;<br>7% |
| quality care while respecting my personal preferences and values |  |  |  |  |  |  |  |
| I was coerced into recommending/administering Covid-19 vaccines against my best clinical judgment (e.g., patient may experience an adverse event, or has experienced an adverse event post-vaccination, Covid-19 or other; patient too young/old to benefit from vaccination, patient experienced Covid-19 and likely has strong natural immunity) | 14/166;<br>8.4% | 17/166;<br>10.2% | 15/166;<br>9% | 13/166;<br>8% | 25/166;<br>15.1% | 72/166;<br>43.4% | 10/166;<br>6% |
| I was accused of undermining Covid-19 public health response/patient care due to my views/decisions about vaccination | 6/166;<br>4% | 7/166;<br>4.2% | 17/166;<br>10.2% | 38/166;<br>23% | 70/166;<br>42.2% | 16/166;<br>10% | 12/166;<br>7.2% |
| I was disciplined for undermining Covid-19 public health response/patient care due to my views/decisions about vaccination | 12/166;<br>7.2% | 9/166;<br>5.4% | 13/166;<br>8% | 22/166;<br>13.3% | 72/166;<br>43.4% | 27/166;<br>16.3% | 11/166;<br>7% |

| Statement (only HCWs who administered vaccines) | Strongly disagree (N; %) | Disagree (N; %) | Neutral (N; %) | Agree (N; %) | Strongly agree (N; %) | N. A | No response |
| --- | --- | --- | --- | --- | --- | --- | --- |
| I am aware that Covid-19 vaccines can cause serious or life-threatening injuries, including death | 0/2;<br>0% | 0/2;<br>0% | 0/2;<br>0% | 1.2;<br>50% | 1/2;<br>50% | 0/2;<br>0% | 0/2;<br>0% |
| I felt coerced to administer Covid-19 vaccines at any point during the vaccination campaign | 0/2;<br>0% | 0/2;<br>0% | 1/2;<br>50% | 0/2;<br>0% | 1/2;<br>50% | 0/2;<br>0% | 0/2;<br>0% |

As well, most respondents (152/166, 92%) responded “no” when asked if they had been encouraged to report adverse events post vaccination if observed in patients. Most (138/166, 83.1%) also responded “no” when asked if they had been trained to report adverse events post vaccination. When asked if they were encouraged or coerced to minimize patients’ hesitancy about vaccines, one third of respondents (57/166; 34%) replied that they were, by, for instance, encouraging patients to trust health officials and official sources (51/166, 31%), telling them that the vaccines were safe and effective (44/166, 27%), dismissing as “misinformation” information that did not align with official sources (38/166, 23%), and denying exemptions (12/166, 7.2%) or off-label prescription (13/166, 8%) requests (Table 8).

Significantly, close to one-fourth of respondents (38/166, 23%) agreed (13/166, 8%) or strongly agreed (25/166, 15.1%) that they had felt coerced to recommend / administer vaccines against their best clinical judgment. As well, most respondents (108/166, 65.1%) agreed (38/166, 23%) or strongly agreed (70/166, 42.2%) that they had been accused of undermining Covid-19 the public health response / patient care due to their reservations about vaccination, and over half (94/166, 57%) agreed (22/166, 13.3%) or strongly agreed (72/166, 43.4%) that they were disciplined for this reason (Table 9). Finally, for two respondents (2/166, 1.22%), job responsibilities included administering vaccines, and one of them (1/2, 50%) reported being reimbursed for the task. Further, when these respondents were asked how they felt upon administering Covid-19 vaccines, both (2/2, 100%) reported feeling uneasy as they were unsure what might happen to vaccine recipients (Table 8). Also, both agreed that they believed that vaccines could cause serious or life-threatening injuries, including death, and one reported that they had felt coerced to administer the vaccine despite their reservations. (Table 9).

## Discussion

In our survey of 166 HCWs in the province of British Columbia, Canada, most of them had 16 or more years of professional experience, were unvaccinated or had not met full vaccination requirements, and had been terminated due to non-compliance with mandates. As well, and regardless of vaccination status, most respondents reported safety concerns with vaccination, yet did not request an exemption due to their experience of high rejection rates by employers. However, most unvaccinated workers reported satisfaction with their vaccination choices, although they also reported significant, negative impacts of workplace mandates on their finances, their mental health, their social and personal relationships, and to a lesser degree, their physical health. In contrast, within the minority of vaccinated respondents, most reported being dissatisfied with their vaccination decisions, as well as having experienced mild to serious post-vaccine adverse events, with more than half of this group reporting having been coerced into taking further doses, under threat of termination, despite these events.

In relation to patient care, a large minority of respondents reported having witnessed underreporting or dismissal by hospital management of adverse events post-vaccination among patients, worse treatment of unvaccinated patients, and concerning changes in practice protocols. About one-third of respondents were encouraged or coerced to minimize patients concerns about, or resistance to, vaccination, by encouraging them to trust health officials and official sources, reassuring them that vaccines were safe and effective, dismissing as misinformation information contradicting official sources, and turning down requests for vaccine exemptions or off-label prescription drugs. Most respondents also reported being accused of undermining patient care due to their reservations about vaccination, and over half reported being disciplined for this reason. As well, most respondents within the very small number of those who administered Covid-19 vaccines reported that they had been coerced into doing so against their best clinical judgment, and that they had felt uncertain about the health impacts of vaccines on recipients. Finally, close to half of respondents reported their intention to leave the healthcare industry altogether.

Our findings in this survey of HCWs in BC differed somewhat from our findings in a similar, earlier survey of HCWs in the province of Ontario (Chaufan et al., 2024). Concerning demographics, respondents in BC tended to be older (33.1% aged 45–54 and 28% aged 55–64) that respondents in Ontario (the largest age group, 29.5%, between 35–44 years). In BC, 10+ years of education/training was the largest category (47%), while Ontario’s largest group reported 0–4 years of education/training (47%), followed by 5–9 years (27%), and then 10+ years (22.4%). The percentage of vaccinated HCWs was lower in BC (13.9%) than in Ontario (25.6%), yet among vaccinated respondents, severe adverse effects were more commonly reported after the first dose in BC (23.1%) than in Ontario (9.2%). As well, in BC the proportion of respondents reporting that employers required additional doses despite adverse effects was greater in BC (54%) than in Ontario (31%).

Mental health and personal impacts also diverged between the provinces. In BC, a larger minority (46%) of respondents than in Ontario (38%) sought counseling. Similarly, BC had a larger percentage of respondents reporting anxiety or depression due to mandates (81.3%) than Ontario (75%). As well, a larger majority (84%) of BC respondents reported that their personal relationships had been negatively affected by mandates as compared to Ontario (73%). Concerning labour relations, a higher percentage of respondents in Ontario (34.2%) than in BC (20%) reported that information about vaccination provided by their employer had not enabled them to make an informed decision, and a higher percentage of BC HCWs (42.2%) reported being subjected to disciplinary measures other than termination compared to Ontario HCWs (22%). These differences likely reflect the fact that mandates in BC were implemented at the level of the province, therefore were harsher and left no room for independent accommodations or decision-making on the part of individual health establishments.

Despite these differences, we identified important similarities in both our Ontario and BC surveys concerning demographics, vaccination decisions, personal and family impacts, and workplace experiences related to Covid-19. In both regions, most respondents were women (67.5% in BC and 82.3% in Ontario), predominantly Caucasian/White (81.3% in BC and 84% in Ontario), born in Canada (74.1% in BC and 77% in Ontario) and married or living with a partner (65% of BC and 72.2% of Ontario respondents). Nursing was the most frequently reported profession (15.1% in BC and 27% in Ontario), with half (50% in BC and 51.2% in Ontario) of respondents reporting 16 or more years of professional experience. Along with our earlier findings, the demographic characteristics of both samples suggest that vaccination mandates in the health sector impacted a largely female and experienced workforce of nursing professionals.

Other studies have also reported greater non-compliance with vaccine mandates among women and nurses, compared to men and other healthcare professionals (Casey et al., 2022; Gogoi et al., 2022; Politis et al., 2023). Among HCWs, women have been more negatively impacted by Covid-19 policies throughout the pandemic (Morgan et al., 2022). Women comprise most of the global healthcare workforce - a phenomenon referred to as the “feminization” of the healthcare workforce (World Health Organization, 2019), so it is expected that women are more likely to experience the effects of policies enacted in healthcare settings. Women also tend to occupy lower status and lower paying healthcare positions (World Health Organization, 2019) and therefore may have less capacity to negotiate workplace policies such as vaccine mandates.

As to vaccination decisions and personal impact of mandates, in both provinces most HCWs reported that workplace mandates were the primary reason for getting vaccinated (78% in BC and 75% of respondents in Ontario), with adverse effects following vaccination widely reported in both provinces (85% in BC and 78% in Ontario). Approximately one-third of respondents in both provinces (31% in BC and 33.3% in Ontario) did not communicate these adverse effects to a doctor, and among those who did, in only a minority of cases (17% in BC and 16% in Ontario) a report was filed. As well, in both provinces most respondents (73% in BC and 66.5% in Ontario) agreed that their income was lower due to vaccination mandates, and most (87.3% in BC and 85% in Ontario) reported feeling unfairly treated by their employers. Furthermore, about one in five respondents in both provinces (Ontario 17% and BC 20%) reported that that their physical health had worsened after mandates, and about one in ten (BC 9% and Ontario 9%) reported that vaccination had led to physical disabilities. The mental health impact of vaccination mandates was also significant in both provinces, with most respondents (86.2% in BC and 83% in Ontario) reporting that job termination had negatively impacted their lives.

In alignment with our finding of high rates of adverse events post-vaccination across provinces, there is growing evidence of adverse events, such as myopericarditis from mRNA vaccines (Buchan et al., 2022; Faksova et al., 2024; Fraiman et al., 2022; Karlstad et al., 2022; Li et al., 2021; Mansanguan et al., 2022; Public Health Agency of Canada, 2021; Yonker et al., 2023; Yun et al., 2024) , and concerns about the lack of reproductive toxicity data (UK.Gov, 2022). The World Council for Health, referencing global reporting data, has raised alarms over an unusually high number of adverse events and recommended recalling Covid-19 vaccines due to concerns over safety, along with evidence for waning, or lack of, effectiveness (World Council for Health, 2022). While overall, the peer reviewed literature on vaccine mandates for HCWs has emphasized the message that Covid-19 vaccines are “safe and effective,” even authors supportive of them have acknowledged adverse events as a risk of vaccine mandates (Bradfield & Giubilini, 2021; Law et al., 2022), since “no vaccine is 100% safe” (Bradfield & Giubilini, 2021), and have granted that by the very nature of imposing a mandate on such a large population, “some will experience these adverse events without freedom of choice” (Law et al., 2022) (page 402). Regarding the negative social, economic and health consequences of vaccine mandates, Bardosh et al. have argued that vaccinate or terminate policies violate the human rights principle of the right to work, creating a negative sequalae for the person affected and their dependents by increasing parental stress, social isolation, and economic deprivation in the family unit (Bardosh et al., 2022). Interestingly, in a study conducted with unions in Canada regarding the issue of workplace vaccine mandates, interviewees described these policies as “internally divisive”, and relayed that “no one thought termination was an appropriate penalty for a worker who refused to be vaccinated” (Braley-Rattai & Savage, 2024) (page 162).

In terms of the impact of mandates on healthcare system sustainability, most respondents in BC (80.1%) and in Ontario (72.4%) reported being terminated due to vaccination noncompliance. In both provinces, most respondents (84% in BC and 82% in Ontario) also reported knowing HCWs who took early retirement due to mandates, and a large minority (44.6% in BC and 42.5% in Ontario) indicated their intention to leave the healthcare industry due to negatives experiences of workplace Covid policies.

Most respondents in both provinces (93.4% in BC and 85% in Ontario) also reported not being offered alternatives to vaccination, and while nearly half in both provinces (44% in BC and 47% in Ontario) requested exemptions – primarily on religious (35% in BC and 30% in Ontario) or medical (23% in BC and 23% in Ontario) grounds – these were largely denied. In both provinces as well, most respondents (91% in BC and 88.5% in Ontario), reported that they had safety concerns regarding Covid-19 vaccines, while more than half (58% in BC and 55.6% in Ontario) had medical concerns.

If our findings indicate a trend in the overall HCWs population, the negative impact of vaccination mandates in the workplace cannot be dismissed as negligible, as claimed by multiple authorities in both provinces, even to this day (Burns, 2023; D’Avino, 2024). The high levels of stress, anxiety, depression, burnout, moral distress and trauma already experienced by HCWs, and exacerbated by the Covid-19 policy response, especially on frontline workers, has been well documented (Bardosh et al., 2022; Benfante et al., 2020; Burrowes et al., 2023; Prasad et al., 2021; Sinsky et al., 2023; Søvold et al., 2021). As a result, HCWs have been leaving their jobs in large numbers – what has been labeled “The Great Resignation” (Linzer et al., 2022). Analysis of US data has also revealed how Covid-19 vaccine mandates for HCWs intensified workforce shortages, causing a reported 6% decrease in the probability of working in the healthcare sector and greater attrition of HCWs (Abouk et al., 2024). According to a survey conducted with BCNU members in May 2021, 76% of nurses reported that their workload increased and over half of emergency room nurses reported an intent to leave the profession (BC Nurses Union, 2021a). Further, many respondents reported deteriorating mental (82%) and physical health (52%) (BC Nurses Union, 2021a). Likewise, a US study reported that nurses were four times more likely to consider leaving their profession due to Covid-19 (Chu et al., 2021). In this context, clearly any policy that results in further workforce reductions will intensify these challenges and negatively impact healthcare system sustainability (Bardosh et al., 2022).

Regarding patient care, most respondents in both provinces observed concerning changes in patient care or procedures following the onset of Covid-19 (67.5% in BC and 76.3% in Ontario), and these percentages increased upon the introduction of vaccination mandates (73% in BC and 76% in Ontario). Most respondents in both provinces (71.1% in BC and 70.1 in Ontario) also reported observing differential treatment of patients based on their vaccination status, with similarly large percentages (70.5% in BC and 69% in Ontario) perceiving an increase in patient harms related to the Covid-19 vaccine. Despite these concerns, only a small minority (5% in BC and 5.1% in Ontario), felt free to express these concerns to their employers, and even fewer (7% in BC and 8% in Ontario) believed their concerns, when expressed, were documented or acted upon. Also, about one fifth of HCWs in BC (23%) and Ontario (29%) reported being encouraged or coerced into recommending or administering vaccines against their clinical judgment however, and a large majority (65% in BC and 64% in Ontario) were accused of undermining Covid-19 public health response and patient care due to their views or decisions about vaccination. Further, around half (57%) of respondents in BC reported being disciplined for raising concerns about vaccination policies (the question was not asked in Ontario). Finally, among the small number of respondents in the sample (2 in BC and 16 in Ontario) who administered Covid vaccines, all or most of them (2 of 2 in BC and 15 of 16 in Ontario) reported they did so despite believing that these could cause serious or life-threatening injuries.

Overall, these findings strongly suggest that vaccination mandates in the workplace have negatively impacted not only the healthcare labour force, but also the quality of patient care. While many authors have maintained that the benefits outweighed the costs, some have highlighted negative impacts on patient care, including a reduction in the amount of patient care provided per day (Gandhi et al., 2024), greater loss of healthcare staff in rural compared to urban settings (Hatch et al., 2023; Yassi et al., 2023), and a decline in morale among HCWs and in quality of patient care (Hatch et al., 2023). Further, according to the BCNU, nurses in the province are experiencing increasing “moral distress” as they witness the negative impacts of a limited workforce on patient care (BC Nurses Union, 2023). Moral distress is described as the tension between ethically appropriate actions and required duties, and institutional constraints and personal values, which if unresolved can manifest as moral injury (Alonso-Prieto et al., 2022). Moral distress creates barriers to patient care, because it leads to HCWs experiencing stress, anxiety, feelings of helplessness, and changes in empathy (Alonso-Prieto et al., 2022). While to the best of our knowledge the concepts of moral distress and moral injury have not been specifically used in relation to the impact of Covid-19 vaccination mandates on the health sector, our findings suggest that HCWs experienced both when recommending or administering vaccines against their own personal convictions.

This study has limitations. First, we relied on a convenience, rather than a random, sample of HCWs. Despite promoting the study through various channels and inviting HCWs of all vaccination statuses to participate, we primarily recruited unvaccinated HCWs. As a result, our findings are not representative or generalizable, since the majority of HCWs in BC were vaccinated – even before mandates were introduced. Nevertheless, our research addresses a gap, since most prior studies have been conducted with predominantly vaccinated HCWs. On the other hand, this lack of representativeness is not unique. For example, in a systematic review on the attitudes of HCWs towards mandatory vaccination policies, the authors reported a “very important finding”, namely, that their review was potentially biased because respondents included in all studies reported high rates of Covid-19 vaccination (Politis et al., 2023). To our knowledge it has not been suggested that studies in which vaccinated HCWs are overrepresented should be dismissed or not used to inform policy.

Other limitations include that our study is cross-sectional, and since we have only conducted descriptive statistical analyses, we were unable to track changes in patterns over time or examine correlations between variables. Nor was our study adequately powered to apply a meaningful multivariate approach able to fully assess the impact of mandates on specific population groups or regions. While such analyses may disprove our conclusions, they could also reinforce them. For example, the observation that most respondents were not only unvaccinated but also middle-class, Canadian born women, could be interpreted as suggesting a disproportionate, negative impact of mandates along gender lines, exacerbating other gender inequities in the healthcare sector (Hennein et al., 2023; Llop-Gironés et al., 2021). The preponderance of middle-class, white women among participants could also be interpreted as indicating that low income or racialized groups - regardless of gender – may have been unable to afford to resist the policy, in word or action, given the significant costs of noncompliance, thus the lack of representation of these groups in our study. Similarly, we could not identify differences in the impact of mandates across provincial regions, although as mentioned earlier, the media attention to emergency room closures in Interior and Northern Health may indicate differential and negative impacts along regional lines.

## Conclusions

In our earlier, similar study in the province of Ontario, we applied the framework developed by the Organization for Economic Cooperation and Development (OECD) in 2021, with its six evaluation criteria that jointly provide standards to “determine the merit or worth of an intervention” - a policy, a strategy, or an activity. To restate these criteria briefly, “relevance” refers to the extent to which a policy is responsive to its intended, direct or indirect, beneficiaries, “coherence” refers to the extent to which a policy is compatible with other policies in a given setting, “effectiveness” refers to the extent to which a policy “has achieved or is expected to achieve its objectives”, “efficiency” refers to the extent to which a policy is cost-effective compared to alternative courses of action within a reasonable timeframe, “impact” refers to the extent to which a policy generates or is expected to generate significant positive or negative, intended or unintended, effects, and finally, “sustainability” refers to whether the benefits of the policy are likely to last (OECD, 2021). At least by OECD criteria, and assuming that the goal of vaccine mandates for HCWs was to promote safer and better quality care rather than increase vaccination rates irrespective of their usefulness, our findings cast doubt on official claims that the policy was successful – as an example of success, Dr. Bonnie Henry’s claim that it supported “the continued provision of essential services” (Office of the Provincial Health Officer, 2022b) (page 4).

By the criterion of “relevance”, intended beneficiaries, whether HCWs, patients, or communities at large, have been harmed by exacerbated staff shortages, intimidating work environments, and health professionals coerced into acting against their best clinical judgment. By the criterion of “coherence”, mandated vaccinations appear at odds with policies aimed at maintaining adequate staffing levels or guaranteeing the protection of patients’ rights, such as informed consent. By the criterion of “effectiveness”, there is no evidence that the policy has improved patient care rather than worsened it. By the criterion of “efficiency”, there is no evidence that the policy has been more cost-effective than alternatives, such as relying on the superiority of naturally acquired immunity over artificial immunity (Altarawneh et al., 2022; Gazit et al., 2022). This is the type of immunity that many HCWs acquired, often working beyond the call of duty in 2020, before vaccines were available, yet to our knowledge, those HCWs were never deemed a threat to patient safety. By the criterion of “impact”, our findings suggest that at least in some settings the overall impact of the policy - on the well-being of HCWs, their families, and their communities - was negative. Finally, by the criterion of “sustainability”, our finding that close to half of our sample of highly trained and experienced HCWs intended to leave the health professions falsifies the claim that mandated vaccinations have not damaged the ability of the health system to provide high quality patient care.

In conclusion, by OECD standards the policy of workplace vaccine mandates in BC has failed. The causes of its failure were predictable and the evidence supporting the prediction was abundant. Future research should address the reasons why, despite multiple and easily identifiable downsides, the policy continued for so long in British Columbia - longer than in any other Canadian province, and well beyond its initial, real or imagined, utility. Researchers and policymakers should also assess the threat that this and comparable policies that involve coercion represent to long-standing bioethical principles, such as informed consent and bodily autonomy. Finally, they should also examine questions of accountability among decision makers in healthcare matters, with a view to guaranteeing better quality care and respect for ethical standards moving forward.

## Author contributions

C.CH designed the project, oversaw the data collection and analysis, and drafted the first version of the manuscript. NH conducted the literature review and made substantive intellectual contributions. NH and RM assisted with data collection and analysis and participated in writing the manuscript. All authors have read and approved the current version.

## Data Availability

All data produced in the present work are contained in the manuscript

## Acknowledgments

CC thanks the many professional and lay organizations, students, trainees, and friends who have afforded spaces of reflection and debate over the past years, and especially her husband Julian Field, for his editorial feedback and support. NH thanks her family and friends for their encouragement and support, and Dr. Chaufan for her mentorship. RM thanks her friends, family, and Dr. Chaufan for their support and guidance. All authors are grateful to the participants for sharing with us their life experiences and making this study possible.

